# What We Learned from COVID-19: From endotheliitis to treatment

**DOI:** 10.1101/2021.07.05.21259790

**Authors:** Adem Dirican, Selin Ildir, Tugce Uzar, Irem Karaman, Sevket Ozkaya

## Abstract

**Objective:** COVID-19 may yield a variety of clinical pictures, differing from pneumonitis to Acute Respiratory Distress Syndrome (ARDS) along with vascular damage in the lung tissue, named as *endotheliitis*. To date, no specific treatment strategy was approved by any authority for the prevention or treatment of COVID-19 in terms of endotheliitis-related comorbidities. Here, we present our experience of COVID-19 by evaluating 11,190 COVID-19 patients with the manifestations of endotheliitis in skin, lung, and brain tissues according to the different phases of COVID-19.

**Methods:** After a retrospective examination, patients were divided into three groups according to their repercussions of vascular distress, which were represented by radiological, histopathological, and clinical findings. (Group A: no or mild pulmonary involvement, Group B: moderate pulmonary involvement with clinical risk of deterioration, Group C: severe pulmonary involvement and respiratory failure). We presented the characteristics and disease course of seven representative and complicated cases which represents the different phases of the disease, and discussed the treatment strategies in each group. The current pathophysiological mechanisms responsible from SARS-CoV-2 infection, COVID-19 related respiratory failure and current treatment strategies were reviewed and discussed in detail.

**Results:** Among 11.190 patients, 9294 patients met the criteria for Group A, and 1376 patients were presented to our clinics with Group B characteristics. Among these patients, 1896 individuals(Group B and Group C) were hospitalized. While 1220 inpatients were hospitalized within the first 10 days after the diagnosis, 676 of them were worsened and hospitalized 10 days after their diagnosis. Among hospitalized patients, 520 of them did not respond to group A and B treatments and developed hypoxemic respiratory failure (Group C) and 146 individuals needed ventilator support and were followed in the intensive care unit, and 43 (2.2%) patients died.

**Conclusion:** Distinctive manifestations in each COVID-19 patient, including non-respiratory conditions in the acute phase and the emerging risk of long-lasting complications, suggest that COVID-19 has an *endotheliitis*-centred thrombo-inflammatory pathophysiology. Endotheliitis can also explain the mechanism behind the respiratory failure in COVID-19, and the difference of COVID-19 related ARDS from ARDS seen in other critical conditions. In addition, use of early corticosteroid in patients with early symptoms and early tocilizumab in ICU helps to reduce mortality and progression of the disease. Endotheliitis-based pathophysiological mechanisms are known to be momentarily changing and difficut to manage due to their risk of sudden aggrevation. Hence, daily evaluation of clinical, laboratory and radiological findings of patients and deciding appropriate pathophysiological treatment would help to reduce the mortality rate of COVID-19.

## Introduction

Novel coronavirus disease 2019 (COVID-19) has resulted in a dramatic pandemic crisis by causing mainly a respiratory disease that can rapidly progress to pneumonia and, in severe cases, to acute respiratory distress syndrome (ARDS) (1). Globally, as of July 15^th^ of 2021, there have been 187,519,798 confirmed cases of COVID-19, including 4,049,372 deaths, reported to World Health Organization (WHO)(2). The heterogeneity of the disease serves a spectrum from asymptomatic cases to respiratory failure. With the experiences from 11.190 COVID-19 patients since March 2020, we observed that distinctive clinical, radiological and histopathological manifestations of each COVID-19 patient, including non-respiratory conditions in the acute phase and the emerging risk of long-lasting complications, suggests an *endotheliitis*-centred thrombo-inflammatory pathophysiology for COVID-19 (3). Therefore, understanding the pathophysiology of SARS-CoV-2 infection, and more significantly the host response against it, is a fundamental tool to develop a personalized treatment for the patient’s need and momentary response. Accordingly, we adopted our treatment strategies depending on both the respiratory and vascular distresses observed in patients. In this paper, we aimed to present the clinical, radiological, and histopathologic manifestations of COVID-19 related endotheliitis, as well as present our treatment categories which were focused on preventing endotheliitis-related consequences in different phases of the disease.

## Methods

### Patient Analysis and Classification

All COVID-19 patients that were diagnosed and treated in Samsun VM Medicalpark Hospital, Turkey, between March 2020 and April 2021 were retrospectively evaluated. Patients who were suspected to be infected by SARS-CoV-2 were confirmed with clinic, laboratory (positive reverse-transcriptase– polymerase-chain-reaction assay of nasopharyngeal swabs or serological IgM/IgG rapid antibody test against SARS-CoV-2) and radiologic (consistent HRCT findings) results and included in the study. Demographic characteristics, presenting symptoms of the patients at the time of admission, radiological images, hospitalization status and the presence of need for respiratory support were retrieved from patient records at the time of admission. Confidentiality of the study participants’ information was maintained throughout the study. The study was performed in accordance with the Helsinki Declaration and approval for this study procedure was obtained from the Istinye University Institutional Review Boards/ethical committees with respect to its scientific content.

### Three Main Groups for Different Phases SARS-CoV-2 Infection

This heterogeneous population of patients were divided into three groups (A,B,C) according to their repercussions of vascular distress, which were represented by radiological, histopathological, and clinical findings. Division into three groups were made according to a combined criteria that was adopted from WHO severity classification and the extent of endotheliitis, which were represented by the clinical symptoms, baseline oxygenations status, radiologic findings (chest x-ray/CT findings), and hemodynamic differences (4). Accordingly, three escalating phases of COVID-19 disease progression with associated signs, symptoms, and potential phase-specific treatments were described as early infection phase(Group A), progressive phase(Group B), and severe and dissipative phase(Group C).

The treatment modalities for each group were adopted from current local guidelines, as well as interventions that were later standardized according to clinical observation of momentary changes by two different pulmonologists (5-7).

#### Group A

Group A included patients who had any of the various signs and symptoms of COVID-19 (e.g., fever, cough, sore throat, headache, muscle pain, diarrhea, loss of taste and smell) but did not have shortness of breath. Group A patients either presented no imaging findings of pneumonia, or they presented with the typical radiological representation of minimal patchy, subpleural, peripheral, perivascular ground-glass opacities (GGO) (Fig. 1-I and 1-II). The placement of GGOs in the early presentation of the disease is compatible with the distribution of microvascular capillaries of lung.

**Figure 1.**
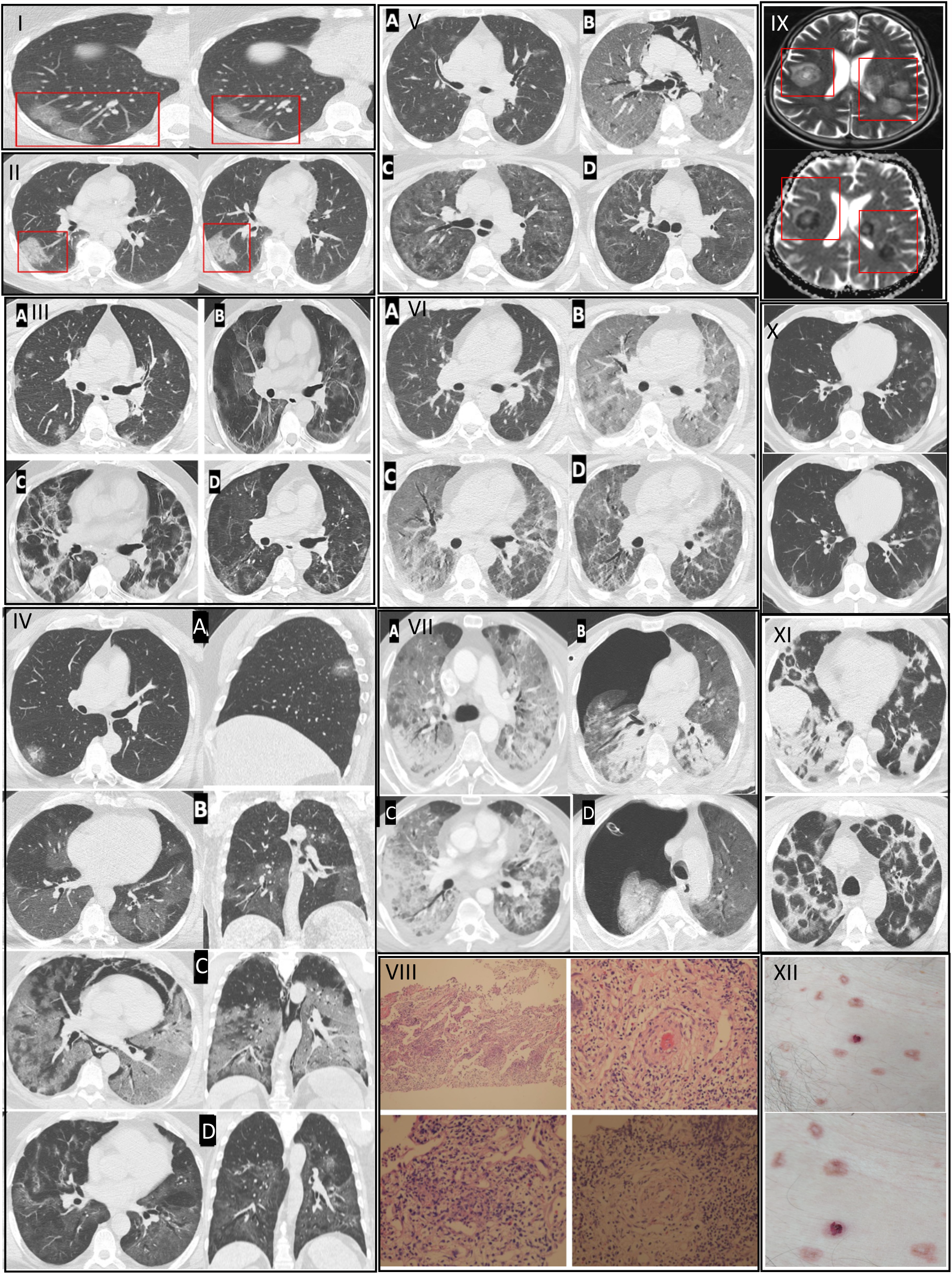
consists of 7 cases with variable stages of COVID-19 and examples of early and severe endotheliitis in brain, pulmonary and skin tissues. Histopathological and radiological findings of pneumonitis changed depending on the phase of the disease (early, progressive, severe and dissipative, respectively), which leads to a divergence in treatment groups. *Examples of patients who were convenient for* ***Group A*** *treatment are depicted in sections* ***I-II***. **Figure 1-I. Case 1**. HRCT images showed patchy, subpleural and peripheral perivascular ground-glass opacities, corresponding the early phase of pneumonitis. She received Group A treatment. GGOs in transaxial images were located in the subpleural area, where the microvascular *endoteheliitis* and endothelial destructions mostly likely occur, due to the interaction with toxic plasma in the capillaries of pulmonary interstitial space. **Figure 1-II. Case 2**. HRCT images showed characteristics of an early phase COVID-19 pneumonitis (end-capillary microangiopathy, explained in Fig.1-I) and perivascular consolidation. It appeared as the continuation of crazy-paving pattern, which was demonstrated by the thickening in inter-lobular and intra-lobular septa. Lung involvement was limited and monofocal. Hence, the patient received Group A treatment. *An example of a patient who was convenient for* ***Group B*** *treatment is depicted in section* ***III***. **Figure 1-III. Case 3**.Transxial HRCT image in the first day of positivity shows bilateral and patchy nodulary GGOs as expected in early phase **(IIIa)**. At the 5th day of positivity, affected pulmonary areas were advanced into scattered consolidations (**IIIb)**. This appearance was noted as the progressive phase of pneumonitis and was considered as representation of clinical deterioation clinically (i.e. dyspnea, respiratory failure). On the 15^th^ day of positivity, fibroreticular consolidations were conspicuous (**IIIc**). The dissipative phase was the healing process, characterized by the resolution in lung parenchyma and residual GGO, observed after 35 days of positivity (**IIId)**. Parenchymal bands, originated from previous fibroreticular proliferation, were also visible (**IIId**). *If the patients have a tendency of severe phase and/or unresponsiveness to Group B therapy*, ***Group C*** *treatment was used. Representative cases of this group are shown in sections* ***IV-V-VI-VII***. *This group of patients also undergo early and progressive phases of COVID-19 but they also showed further deterioration of lung involvement and endothelial damage, therefore, they received the last group of treatment. For the description of early and progressive phases, sections* ***I, II*** *and can be referred*. **Figure 1-IV. Case 4**. Coronal, sagittal and axial planes of HRCT images initially showed small GGO with a local subpleural sparing, particularly around the microvascular area on the first day of positivity (**IVa**). In the 5^th^ day of treatment, increased GGOs were visible in the progressive phase (**IVb**). After 18 days of positivity, pneumomediastinum, Diffuse Alveolar Damage (DAD) and ARDS were seen in the severe phase (**IVc**). Lastly, the dissipative phase was seen, after 30 days of positivity, with residual fibrotic parenchyma (**IVd**). **Figure 1-V. Case 5**. The first HRCT image, on the 2^nd^ day of positivity, showed COVID-19-related bilateral and multifocal nodular GGOs (**Va**). After 10 days of positivity, DAD developed along with ARDS and pneumomediastinum characterized with the severe phase (**Vb**). She recovered and discharged after 30 days of positivity with the dissipative phase (**Vc**). The regression in pulmonary lesions was visible on the HRCT image two months after diagnosis (**Vd)**. **Figure 1-VI. Case 6**. HRCT image showed moderate pneumonia in the early phase (**VIa**). After 12 days of positivity, the severe phase develops with DAD and ARDS (**VIb**). In the 25^th^ day of positivity, recovery was observed in the dissipative phase (**VIc**). The extent of improvement in pulmonary lesions can be noticed in **VId**, which was 40 days after the diagnosis. **Figure 1-VII. Case 7**. First HRCT showed ARDS pattern with dense consolidations (**VIIa**). Pneumothorax developed after two weeks from diagnosis (**VIIb**). In addition to the respiratory failure, hemorrhagical intracranial areas were seen in the T2-weighted MRI. (**VIIc**). He was lost at the 20^th^ day of positivity. *Representative histopathological and radiological images of endotheliitis* ***is seen in section VIII, IX, X, XI***. **Figure 1-VIII** Haematoxylin&Eosin-stained sections from representative areas of lung parenchyma were seen with the mixt-type inflammatory-cell infiltration of lung tissue and exudative capillaritis with thickened microvascular walls. Interstitial and intra-alveolar proliferation of fibroblasts are noted. **Figure 1-IX. Diffusion MRI of** head image of COVID-19 positive patient showing characteristic COVID-19 related *endotheliitis* causing characteristic lesions and intracranial hemorrhage. **Figure 1-X** Early endotheliitis. **Figure 1-XI** Late endotheliitis. **Figure 1-XII** Papulovesicular eruptions as the skin manifestations of COVID-19 related endotheliitis.

If the pulmonary involvement was absent or mild-to-moderate and the patient was suitable for ambulatory treatment, Favipiravir tablet (1600 mg BID for the first day, followed by 600 mg BID for four days, making five days in total), Dexamethasone (0.1-0.2 mg/kg per day), Azithromycin tablet, low-molecular-weight-heparin(LMWH) and acetylsalicylic acid(ASA) therapies were applied for one week.

#### Group B

Patients who showed evidence of lower respiratory disease during clinical assessment (respiratory symptoms) or imaging(findings of pneumonia) but an oxygen saturation(SpO2) ≥94% on room air at sea level, were hospitalized for close follow-up. Patients with comorbidities or special conditions (i.e. age>65, diabetes, cancer, obesity, cardiovascular disease, chronic lung disease, sickle cell disease, chronic kidney disease, being pregnant, cigarette smoking, transplant or immunosuppression recipient) were also hospitalized due to their high risk of severity^4^. Patients who had moderate pulmonary involvement or unresponsive to the Group A treatment in terms of the clinical symptoms, with no evident respiratory failure, but had been indicated for hospitalization were classified as Group B. Group B corresponds to the progressive phase of the disease, which is characterized by multifocal, bilaterally diffused GGOs with poorly circumscribed consolidations scattered in peripheral zones of lungs, along with vascular and intra-lobular septal thickenings called “crazy paving pattern” (Fig.1-IIIb) (7).

For this group, Favipiravir tablet (1600 mg BID for the first day, followed by 600 mg BID for four days, making five days in total), Dexamethasone (0.1-0.2 mg/kg per day) or methylprednisolone (1-2 mg/kg per day), Azithromycin tablet or fluoroquinolone, LMWH, ASA and Famotidine tablet therapies were prescribed. Although the treatment takes one week, we observed that this phase has the peak stage in 10-13 days and may include potential secondary complications. Thus, patients require a close follow-up by serial chest X-rays to establish a baseline to assess the improvement of aeration.

#### Group C

Patients with moderate to severe pulmonary involvement(lung infiltrates >50%.), accompanied by respiratory failure(SpO2 <94% on room air at sea level and, a ratio of arterial partial pressure of oxygen to fraction of inspired oxygen (PaO2/FiO2) <300 mm Hg, respiratory frequency >30 breaths/min), or to the patients who were recalcitrant to standard therapy and had deteriorating clinical status with laboratory (particularly increasing ferritin levels) and radiological findings were classified as Group C. In addition, patients who showed rapid progession(>%50) on CT imaging and who presented with respiratory failure or shock within 24-48 hours also included in this group. This group represents the progressive, peak, dissipative and severe phases of the disease. The radiological findings of Group C were microscopic lacerations and infiltrations in perialveolar vessels which were radiologically appeared as developing pulmonary GGOs, consolidation and diffuse alveolar damage (DAD), which may be accompanied by pneumothorax, pneumomediastinum or intracranial hemorrhage at this stage(Fig. 1-IV-V-VI-VII).

For this severe group of patients, Favipiravir tablet (1600 mg BID for the first day, followed by 600 mg BID for 4 days, continued for 5 or 10 days in total), methylprednisolone (250-1000 mg/day for at least three days), convalescent plasma(once in a day during the first and second days of radiological detoriation, applied for maximum three days), tocilizumab(400 mg QD/daily, applied two times), supported by Piperacilin/Tazobactam, LMWH and famotidine therapies were administered. The effectiveness of this group of treatment was measured by overall mortality ratio.

## Results

A total of 11.190 patients were evaluated. The mean age was 59.2 ±17.3 years and male to female ratio was 5507/5683. Among 11.190 patients, 9294 (83,05%) patients met the criteria for Group A, and 1376 (12.29) patients were presented to our clinics with Group B characteristics. Among these patients, 1896 (16.9%) individuals(Group B and Group C) were hospitalized. While 1220 inpatients were hospitalized within the first 10 days after the diagnosis, 676 of them were worsened and hospitalized 10 days after their diagnosis. Among hospitalized patients, 520 (4.64%) of them did not respond to group A and B treatments and developed hypoxemic respiratory failure (Group C). Among patients who developed hypoxemic respiratory failure, 146 (7.7%) individuals needed ventilator support (non-invasive/invasive mechanical ventilation) and were followed in the intensive care unit, and 43 (2.2%) patients died. As a complication, among 11.190 patients, 30 patients developed either spontaneous pneumothorax, pneumomediastinum or subcutaneous emphysema, and 4 patients died with a 13.3% mortality rate.

Radiologic images and histopathologic samples of representative groups are presented in **Figure 1** which consists of 7 representative and complicated cases with variable stages of COVID-19, togethet with additional examples of findings of early and severe endotheliitis in brain, pulmonary and skin tissues. Representative cases for early phase:Group A (Fig.1-I,II), progressive phase:Group B(Fig.1-III), severe and dissipative phase:Group C(Fig.1-IV,V,VI,VII) were presented and cases were presented in Figure 1.

### Case 1(Figure 1-I)

was a female patient who was in her 50’s and admitted to the hospital with complaints of cough and fever. She was diagnosed with COVID-19 due to PCR positivity. HRCT images showed patchy, subpleural and peripheral perivascular ground-glass opacities, corresponding the early phase of pneumonitis. She received Group A treatment. GGOs in transaxial images were located in the subpleural area, where the microvascular *endoteheliitis* and endothelial destructions mostly likely occur, due to the interaction with toxic plasma in the capillaries of pulmonary interstitial space. She was completely recovered at 14^th^ days at control appointment, after 5 days of treatment with favipiravir.

### Case 2(Figure 1-II)

was a male patient who was 47-52 years old and admitted to the hospital with complaints of fever and chest pain. His COVID-19 PCR test was positive. In addition to the characteristics of an early phase COVID-19 pneumonitis (end-capillary microangiopathy, explained in Fig.1-I) his HRCT images showed perivascular consolidation. It appeared as the continuation of crazy-paving pattern, which was demonstrated by the thickening in inter-lobular and intra-lobular septa. Lung involvement was limited and monofocal. Hence, the patient received Group A treatment. He was completely recovered at 14^th^ days at control appointment, after 5 days of treatment with favipiravir.

### Case 3(Figure 1-III)

was a male patient whose age ranged between 63-68 and admitted to the hospital with complaints of fever and cough. His PCR test resulted positive for SARS-CoV-2. Transaxial HRCT image in the first day of positivity shows bilateral and patchy nodulary GGOs as expected in early phase **(IIIa)**. He received group B treatment and hospitalized for close follow-up. At the 5th day of positivity, affected pulmonary areas were advanced into scattered consolidations (**IIIb)**. This appearance was noted as the progressive phase of pneumonitis and was considered as representation of clinical deterioation clinically (i.e. dyspnea, respiratory failure). On the 15^th^ day of positivity, fibroreticular consolidations were conspicuous (**IIIc**). The dissipative phase was the healing process, characterized by the resolution in lung parenchyma and residual GGO, observed after 35 days of positivity (**IIId)**. Parenchymal bands, originated from previous fibroreticular proliferation, were also visible (**IIId**).

If the patients have a tendency of severe phase and/or unresponsiveness to Group B therapy, **Group C** treatment was used. Representative cases of this group are shown in sections **IV-V-VI-VII**. This group of patients also underwent early and progressive phases of COVID-19 but they showed further deterioration of lung involvement and endothelial damage, therefore, they received the last group of treatment.

### Case 4(Figure 1-IV)

was a male patient whose age ranged between 53-58 and admitted to the hospital with complaints of fever. His PCR test was positive results positive for SARS-CoV-2. Coronal, sagittal and axial planes of HRCT images initially showed small GGO with a local subpleural sparing, particularly around the microvascular area on the first day of positivity (**IVa**). In the 5^th^ day of treatment, increased GGOs were visible in the progressive phase (**IVb**). After 18 days of positivity, pneumomediastinum, Diffuse Alveolar Damage (DAD) and ARDS were seen in the severe phase (**IVc**). Lastly, the dissipative phase was seen, after 30 days of positivity, with residual fibrotic parenchyma (**IVd**).

### Case 5(Figure 1-V)

was a female patient whose aged ranged between 49-53 and admitted to the hospital with complaints of fever and cough. Her COVID-19 PCR test was positive. Her first HRCT, on the 2^nd^ day of positivity, showed COVID-19-related bilateral and multifocal nodular GGOs (**Va**). After 10 days of positivity, DAD developed along with ARDS and pneumomediastinum characterized with the severe phase (**Vb**). She recovered and discharged after 30 days of positivity with the dissipative phase (**Vc**). The regression in pulmonary lesions was visible on the HRCT image two months after diagnosis (**Vd)**.

### Case 6(Figure 1-VI)

was a male patient whose age ranged between 50-55 and admitted to the hospital with complaints of fever and cough. His COVID-19 PCR test was positive. His HRCT showed moderate pneumonia in the early phase (**VIa**). After 12 days of positivity, the severe phase develops with DAD and ARDS (**VIb**). In the 25^th^ day of positivity, recovery was observed in the dissipative phase (**VIc**). The extent of improvement in pulmonary lesions can be noticed in **VId**, which was 40 days after the diagnosis.

### Case 7(Figure 1-VII)

was a male patient whose age ranged between 48-53 and admitted to the hospital with complaint of dyspnea. His COVID-19 PCR test was positive. His first HRCT showed ARDS pattern with dense consolidations (**VIIa**). Pneumothorax developed after two weeks from diagnosis (**VIIb**). In addition to the respiratory failure, hemorrhagical intracranial areas were seen in the T2-weighted MRI. (**VIIc**). He was lost at the 20^th^ day of positivity.

The demonstration of endotheliitis in the biopsy section of lung parenchyma (Fig.1-VIII), radiological images of brain(Fig.1-IX) and lung(Fig.1-X,XI), and skin manifestation(Fig.1-XII) of endotheliitis are also presented in Figure 1.

## Discussion

### Are Laboratory Findings’ Reliable in Determining the COVID-19 Course & Prognosis?

Our classification of patient population and treatment groups was primarily based on the clinical and radiological findings, rather than the laboratory findings. During a year of the pandemic, in our clinics, we observed that even though the laboratory findings may show the degree of pathophysiology during the disease course, it may mislead the clinician when it comes to clinical practice. While C-reactive protein, ferritin, lactate dehydrogenase, erythrocyte sedimentation rate, IL-6 elevation and TNF-alpha represent inflammatory changes; D-dimer, prolonged PTT and troponin elevation show the predisposition to thrombosis and myocardial damage (8,9). Cheng et al. points out that severely affected patients, along with the ones who could not survive the disease, are subject to significantly higher ferritin levels in comparison to the non-severe and survivor groups of patients (10). However, COVID-19 is an acute syndrome of different age and comorbidity groups. When the clinician fails to detect the pre-infectious baseline values of the patient, overtreatment may worsen the situation. In our study group, a patient with 250 mg/L ferritin received Group C treatment, while another with 2550 mg/L received Group A due to the difference in their previous normal values (ferritin increased 10 and <1 times, respectively). The change in the concentrations of inflammatory markers seem to be significantly different in COVID-19 than in typical non-COVID-19-related ARDS, suggesting that COVID-19 features its own unique, poorly understood, yet detrimental, inflammatory profile (11). Therefore, we advise the categorization and treatment strategies to be selected by monitoring radiological findings and the severity of clinical features (such as dyspnea and oxygen saturation).

### Farsighted Evaluation: What are the Causes in Pathogenic Scene After Infection with SARS-CoV-2?

#### Cytokine Storm and Emergence of Toxic Plasma

Pathophysiology of COVID-19 is closely related to cytokine storm, which arises from the consecutive and intricate activation of numerous inflammatory cells that cause excessive and/or unregulated, proinflammatory cytokine release (13). Cytokines force blood plasma to undergo a chemical alteration, revealing toxic and irritant characteristics. Cytokine storm comprises the systemic activation of unstimulated tissue cells, epithelial and endothelial cells in addition to hyperactivation of hematopoietic cells, including B lymphocytes, natural killer (NK) cells, macrophages, dendritic cells, neutrophils and monocytes which provoke the excessive release of proinflammatory cytokines (14). This toxic setting not only causes inflammation but also a damage in various systemic tissues via the signals of pro-apoptosis (15). Main clinical manifestations of cytokine storm appear as fever, progressive dyspnea, tachypnea and elevated inflammatory markers such as IL-6, CRP and ferritin as it is observed in COVID-19 patients (14,15). The uncontrolled production of pro-inflammatory factors (IL-6, IL-8, IL-1β, and GM-CSF), and chemokines (CCL2, CCL3, CCL-5) together with reactive oxygen species (ROS) cause ARDS which leads to pulmonary fibrosis and death. Abnormal nitric oxide metabolism, upregulation of ROS and proteases, downregulation of endothelium-associated antioxidant defense mechanisms, and induction of tissue factor altogether provide a basis for vascular pathology in COVID-19 (3).

Additionally, the systemic inflammatory response against SARS-CoV-2 infection is supported by the circulating mediators found in different organ systems, which are demonstrated in postmortem histopathological analysis to indicate the organ-dependent cytokines (16,17). Fig.1-VIII represents the areas of lung parenchyma with the mixt-type inflammatory-cell infiltration and exudative capillaritis with thickened microvascular walls, in addition to the interstitial and intra-alveolar proliferation of fibroblasts.

#### Vascular Effects: Endotheliitis, Thromboinflammation and Systemic Microangiopathy

Endotheliitis, hypercoagulability and thrombotic microangiopathy are namely the vascular hallmarks of COVIDLJ19 (18,19). These vascular complications should be evaluated separately from ARDS. As a matter of fact, the histopathologic changes observed in several tissue samples might be primarily the result of C3-mediated pathways in thromboinflammation(18). Beigee et al. stated that vascular widespread platelet–fibrin microthrombi was the main pathologic finding in the lung samples of critical COVID-19 patients with severe hypoxemia and minor radiological abnormalities on imaging (16). They also indicated that clinically not all patients with ARDS present DAD. However, the presence of DAD with ARDS contributes worsening of clinical outcomes when compared with those without DAD. Early and late endotheliitis lesions on pulmonary HRCT are seen in Fig.1-X-XI.

Varga et. al were first to demonstrate that irritant plasma, together with the SARS-CoV-2 infection, cause ‘*endotheliitis*’ in microvascular capillary endothelium, which is the primary pathology seen under non-immune, corrosive and irritant conditions (20). Similarly, Zhang et al. demonstrated that COVID-19 infection resembles more of the pathophysiology and phenotype of complement-mediated thrombotic microangiopathies(TMA); rather than sepsis-induced coagulopathy or disseminated intravascular coagulation (21). A common denominator of complement-mediated effects in TMAs is angiocentric inflammation causing endothelial dysfunction; mononuclear and neutrophilic inflammation of microvessels and as a result, microvascular thrombosis, which brings poor prognosis, multiple organ dysfunction syndrome and ARDS(22). Microangiopathies in COVID-19 patients are characterized by anemia, increased lactate dehydrogenase, thrombocytopenia and organ damage (ex. skin lesions, neurological, renal and cardiac dysfunction) (22,23). In our cohort of patients, skin lesions were seen as COVID-19-associated papulovesicular exanthema scattered in trunk and mild pruritus (Fig.1-XII). Trellu et al. indicated that histopathological findings of papulovesicular eruption reveal the signs of *endotheliitis* and microthrombosis in the dermal vessels (24). Unfortunately, patients may die, not from respiratory failure, but due to the vascular coagulopathies (i.e.hemorrhage) in brain, kidneys and heart (Fig.1-IX).

### Fundamentals Behind COVID-19 Related Respiratory Failure

Endothelial dysfunction plays a key role in understanding the multisystemic attack of SARS-CoV-2 infection. Microvascular capillary *endotheliitis* is the primary mechanism that cause clinical detoriation, particularly in those patients with advanced pulmonary involvement (25). The critical point in this manner is to differentiate non-immune *endotheliitis* from immune complex endotheliitis in lungs, and to consider it as the main pathology of COVID-19 since it is not directly mediated by the active antigen-antibody complexes or the virulence of SARS-CoV-2 itself (26,27).

Initially, the irritant action of plasma leads to the thickening of the vessel wall and deceleration in blood flow, which is responsible for the microthrombosis in capillary beds in lungs, which may lead to respiratory failure. Fox et al. demonstrated the microvascular thrombosis and hemorrhage in lungs, as a remarkable contributor of death, in the autopsies of COVID-19 non-survivors (28). They also proved that the cardiovascular damage is “non-immune” by demonstrating cardiac cell necrosis without lymphocytic myocarditis in deceased patients.

The concept of virus-induced pulmonary vasculitis is consistent with a substantial ventilation/perfusion mismatch in COVID-19 based on a right-to-left pulmonary shunt due to a vicious cycle beginning with an increase in respiratory effort and oxygen consumption in inflamed and hyperperfused lungs, failure of hypoxic vasoconstriction, and resulting fatal outcome (3). Therefore, the failure of simple ventilatory support in COVID-19 is commonly observed in patients who are unable to satisfy the oxygen demand due to reduced lung capacity (such as older patients and patients with obesity) and cardiovascular comorbidities.

In the clinical presentation, aggravation of dyspnea and hypoxemia symptoms were attributed to dysfunctional crosstalk between leukocytes and endothelial cells that manifest as vascular immunopathology predominantly confined to the lungs. Eventually, since microvascular walls are prone to damage, the destruction most conveniently occurs as *endotheliitis* at the site of pulmonary interstitial capillaries; with the help of lung elasticity and thin vascular walls, conveys into the perivascular space(29). Remarkably, due to SARS-CoV-2’s endotheliophilic nature, endothelial and epithelial infections appear to be the predominating factors during the course of the disease (3). On the other hand, alveolar-centered infection and the disruption of alveolar epithelial–endothelial barriers contribute to the development of DAD and *pneumonitis*, which manifest as GGOs in alveolar spaces (30,31) (Fig.1-III and VIII). The aforementioned endothelial damage may spread to different systems and become lethal (Fig.1-VII).

Ekanem et al. stated that higher inflammatory markers (ferritin, CRP and fibrinogen), increased fibrosis in HRCT images, and absence of receiving an interleukin-6 inhibitor or convalescent plasma are associated with higher probability of severity and mortality via the spontaneous pneumothorax (SPT) (32). They also suggested that there must be factors uniquely associated with COVID-19 that contribute to the incidence of SPT, since half of the patients were not on a ventilator when the pneumothorax was diagnosed. In Group C (Fig.1-IV-V-VI-VII), we also observed that endothelial damage, along with thromboinflammation, brought increased incidence of pneumothorax secondary to DAD in patients with ARDS. Here, among 11.190 patients, 30 patients developed either SPT, pneumomediastinum or subcutaneous emphysema with a 13.3% mortality rate. The most important reason behind these complications was most likely DAD, which stems from the high transpulmonary pressure and alveolar wall vulnerability, with decreased compliance and increased frailty, resulting in an air leakage into the chest compartments(33). SPT that is observed in severe COVID-19 patients, is thought to be derived from reduced alveolar vessel caliber due to the virus-induced cytolysis, mononuclear immunological response to injury and the small vessel thrombosis at the site of perialveolar area, which should be differentiated from iatrogenic pneumothorax related to mechanical ventilation (34).

### Prevention of Aforementioned Microvascular Pathology and Mortality

Planning an effective therapy for COVID-19 infection is a complex process. According to Mastellos et al. broader pathogenic involvement of C3-mediated pathways in thromboinflammation supports the utilization of complement inhibitors in COVID-19, which result in diminished hyper-inflammation and marked lung function improvement(35). Teuwen et al. suggested that normalization of vascular walls through metabolic interventions might be considered as an additional potential target for the therapy(36). Therefore, until a specific antiviral is discovered against SARS-CoV-2, convalescent plasma therapy and immunomodulators play a significant role to control the consequences of SARS-CoV-2 infection (i.e. cytokine storm), to reduce inflammatory cell infiltration in lungs and to prevent fatal course in severe patients by reducing the likelihood of the Systemic Inflammatory Response Syndrome (37).

The latest COVID-19 treatment guideline published by the Turkish Ministry of Health included the favipiravir, ASA, famotidine, LMWH, dexamethasone, tocilizumab, which were shown to be safe and in vitro effective against SARS-CoV-2 (5,6). Based on indirect evidence from different clinical trials, convalescent plasma therapy and pulse steroid therapy were also suggested for severe patients (38). The required interventions were made according to clinical severity of patients, vitals, time from symptom onset and radiological images.

***Convalescent plasma*** therapy plays a critical role in neutralizing the plasma and diminishing its corrosiveness. Convalescent plasma not only demonstrates an antibody response but also denotes immunomodulatory, anti-cytokine and proinflammatory effects, which appear as key factors to minimize disease severity and mortality in COVID-19 cases(38). Gomez-Pastora et al. explain this correlation by the phenomena of pro-inflammatory and anti-inflammatory cytokine activation, as a result of macrophage associated hyperferritinemia(39). They stated that ferritin plays an active role as a pathogenic mediator in COVID-19 and the therapeutic use of plasma is beneficial to reduce ferritin and cytokine levels in the body. Our experience with convalescent plasma showed rapid and positive results against the symptoms of dyspnea, hypoxemia, fever and radiologically seen infiltrations, which was demonstrated **in Case 4-7**.

***Antiviral drugs*** are being used to decrease the viral load. Correspondingly, Favipiravir was the drug of choice that is recommended by Turkish Republic of Health Ministry guidelines (5,6). However, it should be noted that antiviral therapy fails to prevent pulmonary involvement, which is the result of inflammatory process rather than the effect of SARS-CoV-2 infection itself (40).

***Systemic corticosteroid drugs*** (dexamethasone and methylprednisolone) are the only effective therapeutic agents to repair non-immune capillary microvascular *endotheliitis*, hence, advised to be used even in the presence of minimal ground-glass opacities(41). In our clinical experience, to benefit the best of steroids, steroids should be utilized in the early phase, rather than the progressive phase (approximately the second week of infection). Minimally distributed GGOs may easily progress to severe ARDS, in the absence of steroid treatment (**Fig.1-VIIa-VIb**). In a multi-centered study conducted by RECOVERY Collaborative Group, mortality ratio in patients, who receive oxygen support and dexamethasone, found to be lower than the control group, especially if the patients are receiving invasive mechanical ventilation (29.3% vs. 41.4%; rate ratio, 0.64; 95% CI, 0.51 to 0.81) (42). Although the role of corticosteroids in COVID-19 has been well-recognized in the therapeutic algorithm, the right timing, dosage and duration of corticosteroid use is still unknown. Pinna et al. suggested that although the early use of corticosteroids might facilitate the viral replication in the upper airways, late administration fails to prevent the alveolar damage(43). It has been suggested that early initiation of low-dose methylprednisolone therapy (with up to 40mg/day for less than 10 days) provides clinical, radiological improvement in patients with active disease state of COVID-19, by reducing the immune cascade and progression into cytokine without any adverse effect (44-46).

*Immunomodulatory therapies* help to diminish the cytokine response of the body. Tocilizumab therapy, a monoclonal IL-6 antagonist, reduced the likelihood of progression to the composite outcome of mechanical ventilation or death (47). Capra et al. have demonstrated that tocilizumab can be used as an immunomodulatory drug of choice in case of the severe COVID-19 to reduce mortality, diminish oxygen intake and treat lung opacities as well (48). Findings by Gupta et al. also suppoerted the early use of(within first 2 days of ICU) tocilizumab to reduce the in-hospital mortality among critically ill patients with COVID-19 (49).

***LMWH and ASA*** are well-known to prevent the formation of microvascular thrombosis and cure hypoxemia, thus can be used as supportive treatment against cardiovascular complications of COVID-19 (50). However, before the use of ASA, patient history should be questioned in terms of renal failure, gastrointestinal system disease, cardiovascular disease. Local current guidelines recommended the use of NSAIDs, especially in the first 5-10 days; and steroid treatment to be started in the early period in patients who worsen after theoretical viral clearance is completed in the first 5-10 days (5).

### Limitation

We acknowledge that there are confounding factors related to management of COVID-19 patients due to lack of a standardized guideline for treatment of each and every patient. However, while the in-hospital mortality was reported to be up to 25% in different prospective trials, we believe that our standardized treatment approach, which has a result of 2.2% in-hospital mortality, represent the success of the personalized management of each COVID-19 patient in a single center (50). As the COVID-19 pandemic continues, our current strategy represents a snapshot that would most probably change drastically over time.

As a limitation, the design of the current study does not allow for further analysis of the COVID-19 patients in terms of the change in their laboratory, clinical and radiological parameters and investigation of the effects of different SARS-CoV-2 variants(such as Alpha, B.1.1.7, Beta B.1.351, or Delta B.1.617.2) in the clinical presentation of the patients. The observational methodology suggested may be a source of bias that could lead to wrong conclusions on the effectiveness of treatments and the clinical representation of the underlying pathophysiology. Provided treatment strategies are implemented to all patients regardless of their SARS-CoV-2 variant types. No corruptions from Turkey’s Ministry of Health guidelines are enforced to the patients in terms of new experimental drugs and/or biological agents.

We would like to declare that, our study did not interfere any patient’s right to recieve treatment by adressing a control group in a pandemic situation. Here, we also did not aim to demonstrate the effectiveness of the therapy in the means of laboratory data, however we aimed to present the representations of the required interventions in the each group of patients, by preventing the progression of endotheliitis. Therefore, we used the current literature to support our clinical observation and own perspective. In this end of the study, our aim was to declare our own point of view by using our clinical experience, clinical representation of endotheliitis and the current literature. We would like to approach to the vascular distress phenomenon as a clinical parameter that can be used practically in the clinics, both by recognizing from radiological images and clinical findings of the patients

## Conclusion

SARS-CoV-2 infection is a multisystemic disease which courses rapidly with respiratory failure and complications secondary to vascular alterations (i.e. microvascular thrombosis, *endotheliitis*, and cytokine induced plasma toxicity). Early detection of radiological detoriation before laboratory findings, via monitoring chest X-rays daily, and planning personalized treatments constitute a crucial and life-saving maneuver in the treatment of COVID-19. Our group suggests that an important key to success relies on how closely the clinicians follow patients from diagnosis to treatment, including the whole course of the disease from outpatient clinic to ICU, in order to differentiate instant clinical deviations from previous general status

Distinctive manifestations in each COVID-19 patient, including non-respiratory conditions in the acute phase and the emerging risk of long-lasting complications, suggest that COVID-19 has an *endotheliitis*-centred thrombo-inflammatory pathophysiology. Potential pathophysiological mechanisms contributing to endotheliitis includes cytokine storm and toxic plasma, thromboinflammation and systemic microangiopathy. Endotheliitis can also explain the mechanism behind the respiratory failure in COVID-19, and the difference of COVID-19 related ARDS from ARDS seen in other critical conditions. In our observations, utilization of early dexamethasone in Group A prevented the progression of the COVID-19 into more severe form. In addition, use of early steroid in Group A and early tocilizumab in group C helps to reduce mortality and progression of the disease. Endotheliitis-based pathophysiological mechanisms are known to be momentarily changing and difficut to manage due to their risk of sudden aggrevation. Hence, daily evaluation of patients and deciding appropriate pathophysiological treatment for the momentary changes in clinical, laboratory and radiological findings would help to reduce the mortality rate of this novel virus. The collaboration of scientists and clinicians around the world is required in order to develop novel prognostic biomarkers and establish precise predictive thresholds for known biomarkers to foresee the severity for COVID-19 pneumonitis that is characterized by vasculopathies and wide range of immune derangements.

**Table 1.**
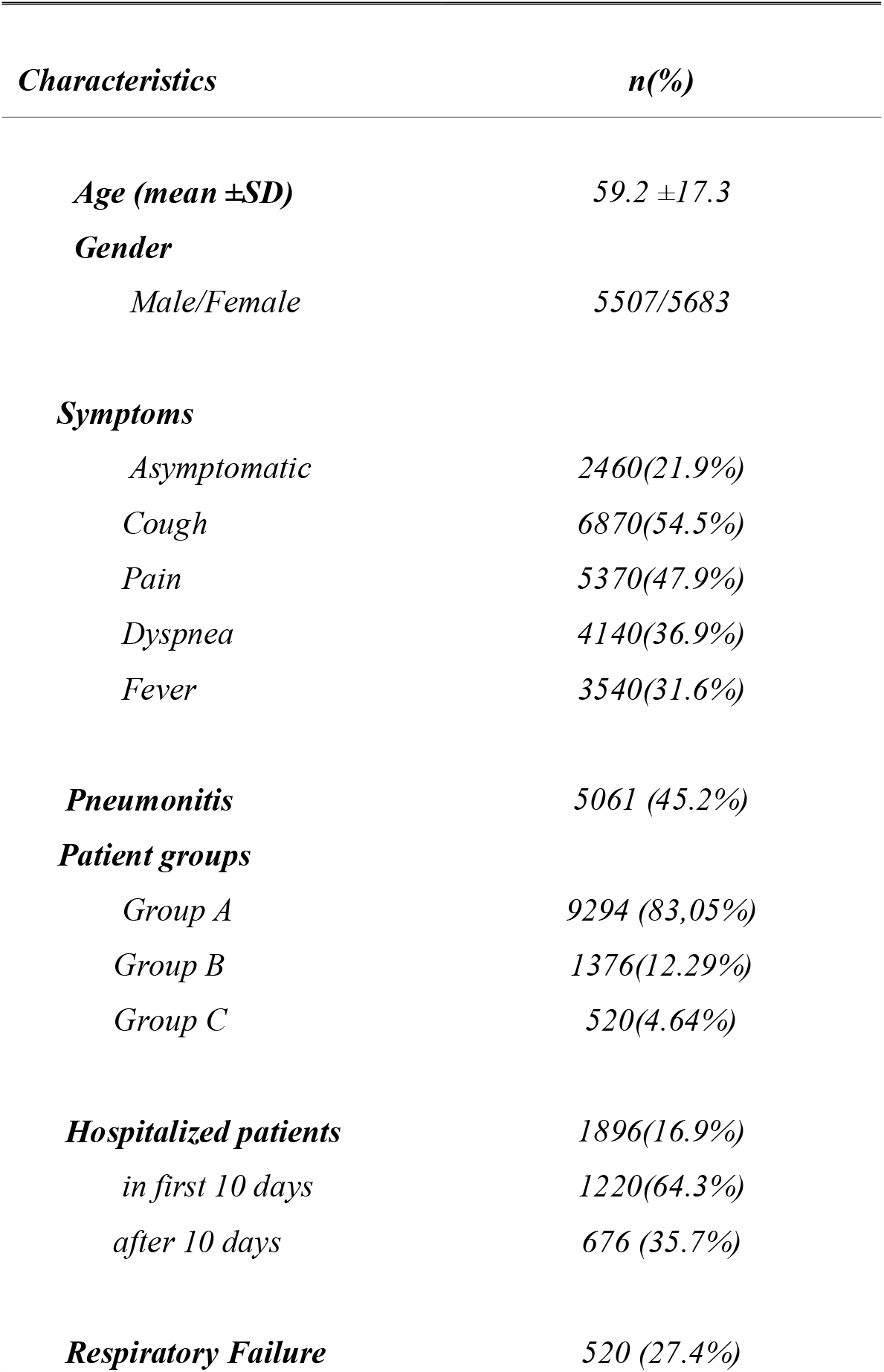
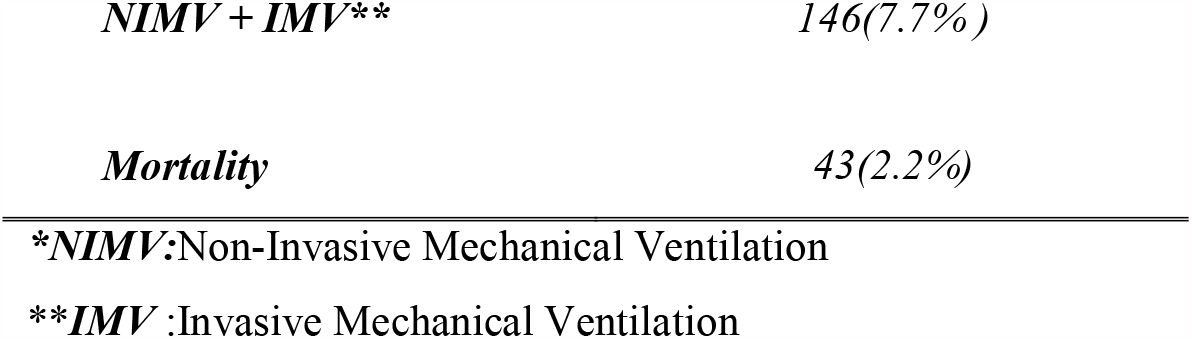
The Characteristics of Patients with COVID-19.

## Data Availability

The research article data used to support the findings of this study are available from the corresponding author upon request.

## Ethics Statement

The patients signed written informed consents to be able to provide data in this study. The study was performed in accordance with the Helsinki Declaration and approval for this study procedure was obtained from the Istinye University Institutional Review Boards/ethical committees with respect to its scientific content.

## Authors’ Contribution

AD and SO diagnosed, treated the patients and designed the analysis, SI, TU and IK collected the data, analysis tools and wrote the paper.

